# Time from onset to aneurysm securing is associated with rebleeding in patients with aneurysmal subarachnoid hemorrhage

**DOI:** 10.1101/2025.01.02.25319926

**Authors:** Emile Daurel, Arthur Le Gall, Claire Charamel-Lenain, Alexandre Ledos, François Eugène, Yoann Launey

## Abstract

Rebleeding is a major complication in patients admitted for aneurysmal subarachnoid hemorrhage. The optimal timing for aneurysm securing after the initial headache remains debated. The objective of this study was to evaluate the previously established predictive factors for rebleeding in a contemporary retrospective cohort (2020-2022, Rennes University Hospital). Among the 226 patients included in the analysis, 33 (15%) experienced rebleeding. The mean time-to-event (rebleeding or securing) was significantly shorter in patients without rebleeding compared with patients with rebleeding (36 ± 3 *vs.* 63 ± 9 hours, respectively, p<0.001). For each additional hour of delay since the first symptoms, the odds of rebleeding increased by 0.7% (OR = 1.007 [1.001-1.013], p=0.026). This association remains significant in multivariate analysis. Despite its limitations, this study highlights the importance of early management, taking into account the time elapsed since the initial headache.

## Introduction

Rebleeding is a major complication in patients admitted for aneurysmal subarachnoid hemorrhage (aSAH), associated with high mortality and morbidity.

The factors that have been identified as predictors of rebleeding include high systolic blood pressure, higher grades of the Word Federation of Neurosurgical Societies (WFNS) scale, intracerebral or intraventricular hematoma, aneurysm size and aneurysm located on the posterior circulation territory (1–11). Also, a particular attention has been given to the time elapsed since aneurysmal rupture, especially with advancements in surgical clipping and endovascular coiling techniques.

Currently, since early treatment (within 48-72 hours of aSAH) has been associated with favorable outcomes (12–21), the European and American international societies recommend that the aneurysm should be treated as early as logistically and technically possible and, if possible, at least within 72 hours after onset of first symptoms (22) or within 24 hours of onset (23), respectively. However, it appears that the optimal timing for securing a ruptured aneurysm remains debated (24–27).

We hypothesize that improved stratification of rebleeding risk, based on established predictors, could optimize the timing of therapeutic decision-making. The objective of this study was to evaluate the previously established predictive factors for rebleeding in a contemporary retrospective cohort.

## Methods

### Patients

In this retrospective single-center cohort study, we included adult patients (aged 18 years or older) admitted to Rennes University Hospital between January 2020 and December 2022, for aSAH, confirmed by head computed tomography (CT) scan.

We collected data on patient demographics (age, gender, body mass index, medical history, tobacco and alcohol use), initial clinical and radiological assessments (systolic blood pressure, WFNS and Fisher grades, presence of intracerebral or intraventricular hematomas, aneurysm location and size), treatment modality (endovascular or neurosurgical), and clinical course (occurrence of hydrocephalus, vasospasm, neurogenic cardiomyopathy, and seizures). Vasospasm was evaluated through a review of medical records. A vasospasm was defined as a diagnosis made by a specialized neuroradiologist based on findings from a head CT scan with perfusion sequencing, followed by therapeutic intervention, such as angioplasty or hemodynamic augmentation. Neurogenic cardiomyopathy was defined as hemodynamic failure, not explained by another cause of hypotension, and confirmed by cardiac ultrasound.

We retrospectively identified the date and time of the initial headache from medical records. In case of neurological deterioration, rebleeding was considered the most likely cause, unless another cause was clearly identified (*e.g.*, seizures or hydrocephalus).

### Outcomes

The primary outcome – the rebleeding – was defined as a new deterioration in the neurological examination occurring after headache or ictus onset and confirmed by a specialized neuroradiologist on a head CT-scanner.

The secondary outcomes were short- and medium-term prognoses, assessed using the Glasgow Outcome Scale (GOS) at 1 and 6 months, respectively.

### Statistical analysis

The population was divided into two groups, the patients with (RB+) and without (RB-) rebleeding. Population characteristics were compared between RB+ and RB-patients. Continuous variables were analyzed using the Student’s t-test, ordinal categorical variables with the Wilcoxon rank-sum test, and nominal categorical variables the Chi-squared test, as appropriate.

Given that the study had less than 10% missing data overall, and less than 1% for the primary outcome, missing values were handled using complete-case analysis.

We performed an unadjusted and adjusted generalized linear regression to investigate the association between the known predictors of rebleeding (systolic blood pressure at admission, WFNS score, posterior location of aneurysm, intraventricular hemorrhage, intracerebral hematoma, aneurysm size, time from the first symptom to event) and the primary outcome. To account for competing risks, data were right censored after the occurrence of an event. The end of the exposure period was aneurysm securing. Model selection was conducted using a stepwise approach, combining backward and forward elimination, to minimize the Akaike Information Criterion (AIC).

A comprehensive analysis was performed to examine the various timing aspects of patient care. To account for non-linearity between time-to-event and the event, a natural spline polynomial regression model was employed. Given the non-normal distribution of the time-to-event variable, a bootstrap procedure was applied to calculate confidence intervals.

A p-value ≤ 0.05 was considered statistically significant.

### Data management and ethics

The study complied with the French reference methodology MR-004 and we obtained the approval from the local ethics committee (decision n°24.82) (28).

## Results

### Descriptive statistics

*Figure 1* and *Table 1* present the study flowchart and the baseline characteristics of the population, respectively.

**Figure 1.**
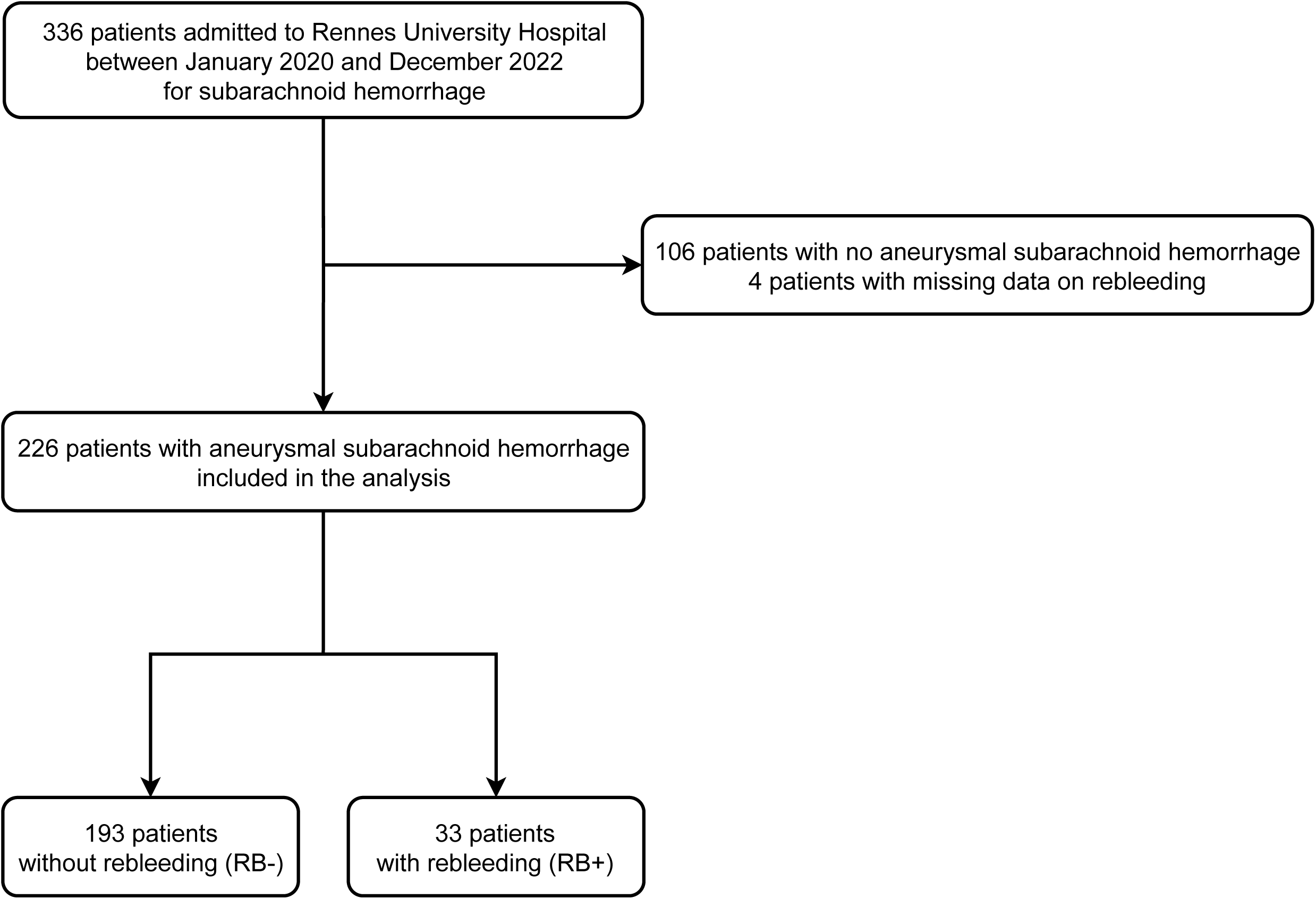
Study flowchart.

**Table 1.** Characteristics of the population. BMI: Body Mass Index; GCS: Glasgow Coma Scale; IQR: Interquartile Range; RB-: patients without rebleeding; RB+: patients with rebleeding; SBP: Systolic Blood Pressure; SD: Standard Deviation; WFNS: World Federation of Neurosurgical Societies.

|  | All (n=226) | RB- (n=193) | RB+ (n=33) | p-value |
| --- | --- | --- | --- | --- |
| **Demographics** |  |  |  |  |
| Age, mean ± SD (years) | 55 ± 14 | 55 ± 15 | 54 ± 13 | 0.725 |
| Gender |  |  |  | 0.462 |
| Female | 159 (70%) | 134 (69%) | 25 (76%) |  |
| Male | 67 (30%) | 59 (31%) | 8 (24%) |  |
| BMI, mean ± SD (kg.m<sup>-2</sup>) | 24.5 ± 4.4 | 24.2 ± 4.2 | 26.2 ± 5.5 | 0.051 |
| **Prior conditions** |  |  |  |  |
| Arterial hypertension | 69 (31%) | 59 (31%) | 10 (30%) | 0.961 |
| Diabetes mellitus | 7 (3%) | 4 (2%) | 3 (9%) | 0.032 |
| Tobacco use | 128 (57%) | 112 (58%) | 16 (48%) | 0.265 |
| Alcohol abuse | 32 (14%) | 29 (15%) | 3 (9%) | 0.360 |
| **Severity scores** |  |  |  |  |
| GCS, median [IQR] | 15 [9-15] | 15 [9-15] | 15 [9-15] | 0.874 |
| WFNS |  |  |  | 0.437 |
| I | 114 (50%) | 98 (51%) | 16 (48%) |  |
| II | 33 (14%) | 31 (16%) | 2 (6%) |  |
| III | 3 (1%) | 2 (1%) | 1 (3%) |  |
| IV | 41 (18%) | 33 (17%) | 8 (24%) |  |
| V | 35 (15%) | 29 (15%) | 6 (18%) |  |
| Fisher |  |  |  | 0.980 |
| 1 | 26 (12%) | 22 (11%) | 4 (12%) |  |
| 2 | 15 (6%) | 13 (7%) | 2 (6%) |  |
| 3 | 36 (16%) | 30 (16%) | 6 (18%) |  |
| 4 | 149 (66%) | 128 (66%) | 21 (64%) |  |
| SBP, mean ± SD (mmHg) | 144 ± 27 | 144 ± 27 | 144 ± 25 | 0.998 |
| **Imaging** |  |  |  |  |
| Aneurysm location |  |  |  | 0.290 |
| Anterior cerebral artery | 16 (7%) | 15 (8%) | 1 (3%) |  |
| Middle cerebral artery | 58 (26%) | 53 (27%) | 5 (15%) |  |
| Anterior communicating | 73 (32%) | 61 (32%) | 12 (36%) |  |
| Internal carotid artery | 55 (24%) | 46 (24%) | 9 (27%) |  |
| Posterior circulation | 24 (10%) | 18 (9%) | 6 (18%) |  |
| Aneurysm size, mean ± SD (mm) | 6.5 ± 3.8 | 6.4 ± 3.6 | 7.5 ± 4.8 | 0.234 |
| Intracerebral hematoma | 63 (28%) | 51 (26%) | 12 (36%) | 0.239 |
| Intraventricular hematoma | 129 (57%) | 113 (59%) | 16 (48%) | 0.280 |
| **Aneurysm treatment** |  |  |  | 0.324 |
| Endovascular | 192 (85%) | 163 (84%) | 29 (88%) |  |
| Neurosurgical | 27 (12%) | 25 (13%) | 2 (6%) |  |
| **Complications** |  |  |  |  |
| Hydrocephalus | 92 (41%) | 77 (40%) | 15 (45%) | 0.548 |
| Vasospasm | 109 (48%) | 89 (46%) | 20 (61%) | 0.124 |
| Neurogenic cardiomyopathy | 17 (8%) | 15 (8%) | 2 (6%) | 0.730 |
| Seizures | 47 (21%) | 39 (20%) | 8 (24%) | 0.598 |

Between January 2020 and December 2022, 336 patients were admitted to Rennes University Hospital for subarachnoid hemorrhage. Among these patients, 226 were included in the analysis after excluding 106 patients with non-aneurysmal subarachnoid hemorrhage and 4 cases with missing data on rebleeding.

Most patients were women (70%), aged 40–60 years (mean ± standard deviation (SD): age=55 ± 14 years), smokers (57%), non-obese (mean ± SD: BMI=24.5 ± 4.4 kg.m^-2^) and non-diabetic (3%). Most patients underwent endovascular treatment (85%).

Rebleeding occurred in 33 patients (15%). No statistically significant differences were observed between the RB- and RB+ groups for the main risk factors for rebleeding identified in the literature.

Systolic blood pressure at admission was similar between RB+ and RB-patients (p=0.998). Severity scores such as WFNS and Fisher grades and aneurysm location were comparable between both groups (p=0.437, p=0.980, and p=0.290, respectively). Aneurysm size did not significantly differ (p=0.234).

The incidence of complications (hydrocephalus, vasospasm, neurogenic cardiomyopathy, or seizures) was also comparable between RB+ and RB-patients (p=0.548, p=0.124, p=0.730, and p=0.598, respectively).

At 1 and 6 months, prognosis of RB- and RB+ patients was not statistically different. However, a trend toward different GOS scores was observed at 6 months between RB- and RB+ patients (p=0.052). In the subgroup analysis of GOS 5 scores, we observed a higher incidence in the RB-group compared to the RB+ group (p = 0.03).

### Association with outcomes

*Table 2* summarizes the results for the primary and secondary outcomes. In univariate analysis, among the predictors for rebleeding described in the literature, only the time-to-event was associated with the risk of rebleeding. For each additional hour of delay since the first symptoms, the odds of rebleeding increased by 0.7% (OR = 1.007 [1.001-1.013], p=0.026). In multivariate analysis, we observed that this association remained significant (p=0.008). We also observed an independent linear relationship between WFNS score and rebleeding risk (OR = 2.444 [1.042-5.749], p=0.039).

**Table 2:** Outcomes. GOS: Glasgow Outcome Scale; RB-: patients without rebleeding; RB+: patients with rebleeding; SD: Standard Deviation.

|  |  | All (n=226) | RB- (n=193) | RB+ (n=33) | p-value |
| --- | --- | --- | --- | --- | --- |
| **Times to** |  |  |  |  |  |
| Event, mean ± SD (hours) | | 35 ± 18 | 36 ± 3 | 63 ± 9 | <0.001 |
| Admission, mean ± SD (hours) | | 28 ± 14 | 18 ± 2 | 60 ± 9 | <0.001 |
| Aneurysm securing, mean ± SD (hours) | | 43 ± 22 | 36 ± 3 | 87 ± 11 | <0.001 |
| **Prognosis** |  |  |  |  |  |
| GOS 1 month |  |  |  |  | 0.519 |
|  | 1 | 34 (15%) | 27 (14%) | 7 (21%) |  |
|  | 2 | 10 (4%) | 10 (5%) | 0 (0%) |  |
|  | 3 | 52 (23%) | 44 (23%) | 8 (24%) |  |
|  | 4 | 68 (30%) | 58 (30%) | 10 (30%) |  |
|  | 5 | 61 (27%) | 54 (28%) | 7 (21%) |  |
| GOS 6 months |  |  |  |  | 0.052 |
|  | 1 | 40 (18%) | 31 (16%) | 9 (27%) |  |
|  | 2 | 1 (0%) | 1 (1%) | 0 (0%) |  |
|  | 3 | 37 (16%) | 34 (18%) | 3 (9%) |  |
|  | 4 | 58 (26%) | 45 (23%) | 13 (39%) |  |
|  | 5 | 87 (39%) | 80 (42%) | 7 (21%) |  |

### Timing analysis

The mean time-to-event (defined as the time to rebleeding or the time to securing) was significantly shorter in RB-patients compared with RB+ patients (36 ± 3 *vs.* 63 ± 9 hours, respectively, p<0.001). Notably, two-third of rebleeding events occurred more than 24 hours after symptom onset. Among the 114 (50%) patients whose aneurysms were secured within 24 hours, 10 (9%) rebled prior to securing, compared with 23 (21%) in the 112 treated patients after 24 hours (p=0.021).

Time to hospital admission was also significantly shorter in RB-patients compared with RB+ patients (18 ± 2 *vs.* 60 ± 9 hours, respectively, p<0.001).

Although time from admission to treatment was slightly shorter in RB-patients (18 ± 1 *vs.* 19 ± 1hours, p<0.001), this 1-hour difference is unlikely to be clinically relevant.

Among patients who experienced rebleeding after hospital admission (n=11), we observed a median delay of 4 hours [IQR 3–9] from admission to rebleeding. However, the overall time from onset to rebleeding was shorter than 24 hours, with a median of 7 hours [IQR 5–15.5].

### Probability of rebleeding over time

*Figure 2* illustrates the probability of rebleeding as a function of time since the initial headache.

**Figure 2.**
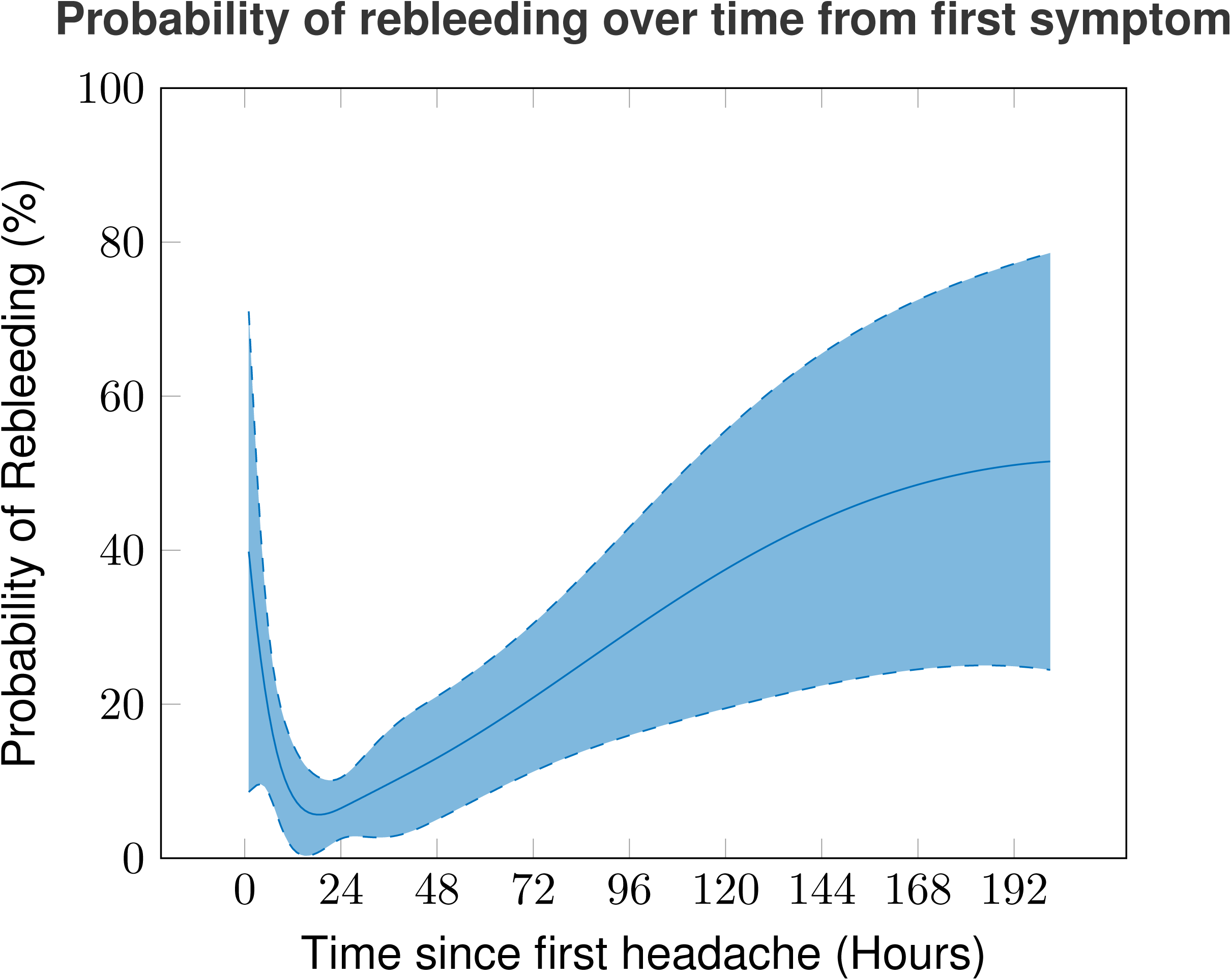
Probability of rebleeding over time from first symptoms.

Applying these findings to a hypothetical population with admission and securing timings similar to our study population, we would have observed a 41 ± 1% increase in the risk of rebleeding at the time of patient admission and a 50 ± 1% increase at the time of aneurysm securing in RB+ patients compared to RB-patients (p<0.001).

## Discussion

This retrospective monocentric study demonstrated that the time elapsed since the onset of the first symptoms of aneurysmal subarachnoid hemorrhage was independently associated with the risk of rebleeding. Additionally, among the factors previously identified as linked to the risk of rebleeding, our cohort analysis confirmed that WFNS score was a reliable independent predictor.

The timing of aneurysm securing after the initial headache has long been debated. Only one prospective randomized trial, conducted in the pre-embolization era, attempted to identify the optimal timing for surgery in patients with ruptured aneurysms in the anterior circulation who were in good conditions (Hunt and Hess grades I to III) (29). Öhman and Heiskanen (1989) demonstrated that acute surgery (0-3 days from aSAH) resulted in lower mortality or morbidity rates at 3 months compared with intermediate (4-7 days) or late (>7 days) surgeries.

Subsequent studies, often retrospective, attempted to confirm these findings, but definitions of “early” and “late” treatment varied across studies. Dorhout Mees (13), in a post-hoc analysis of the International Subarachnoid Aneurysm Trial (ISAT), assessed outcomes at 2 months and 1 year for 2106 patients (>90% WFNS grades I to III), dividing them into 4 categories: treatment within 2 days after the hemorrhage, on days 3-4, on days 5-10, and after 10 days (30). The authors concluded that aneurysm treatment beyond day 10 was associated with worse outcomes, regardless of treatment modality. In a systematic literature review, de Gans *et al.* (12), suggested that both early (days 0-3) and intermediate (days 4-7) surgical treatment improved outcomes in aSAH patients who were in good clinical conditions at admission.

These findings, supporting early treatment, have also been confirmed in patients with poor conditions. A meta-analysis by Zhao *et al.* (21) showed that early treatment, within 48 hours of aSAH, was associated with higher rates of favorable outcomes (61% *vs.* 40%, p<0.01) for poor-grade aSAH.

Modern research has shifted to comparing ultra-early treatment (within 24 hours of aSAH onset) with early treatment. A meta-analysis of 16 studies found that endovascular treatment within 1 day was associated with better outcomes compared with treatment after 1 day, but not significantly better than treatment within 1-3 days (26). Similarly, Luo *et al.* (20) demonstrated that poor-grade aSAH patients had improved clinical outcomes with ultra-early coiling compared to coiling performed after 24 hours (58.1% *vs.* 21.4%, p=0.028).

This 0.7% per hour increase in the risk of rebleeding, though seemingly modest, aligns with and supports the latest 2023 guidelines of the American Heart Association/American Stroke Association. These guidelines recommend that “treatment should be performed as early as feasible after presentation, preferably within 24 hours of onset” (23). Assuming a linear increase in the risk of rebleeding during the first 24 hours, the overall risk of rebleeding at day 1 is approximately 17%. A literature review on ultra-early rebleeding suggests that studies reporting rebleeding rates of about 4% within the first day may have underestimated this risk by failing to capture very early rebleeding events (31). The estimated risk of rebleeding within the first 24 hours ranges from 9% to 17%. This aligns well with our study, which evaluated the time elapsed since the initial headache. Our findings strongly advocate of securing aneurysms as early as possible.

Although the 1 and 6-month prognoses of RB- and RB+ patients were not statistically different, there was a trend toward a higher rate of GOS 5 scores at 6 months in RB-patients, providing additional support for early aneurysm treatment. Using the GOS-Extended (scored 1-8) as a prognostic tool might have yielded statistically significant results.

The limitations of our study include its retrospective nature and the limited number of events, which may affect the robustness of the multivariate analysis.

Also, our definition of rebleeding could be subject to debate. We adopted a pragmatic and clinically oriented definition that may lack definitive confirmation. Specifically, we reviewed medical records to determine the first recorded onset of headache, even if this symptom had not yet been identified as linked to a subarachnoid hemorrhage. We interpreted rebleeding as the subsequent neurological deterioration or recurrence of headache that ultimately led to a diagnosis of aSAH, confirmed by a CT scan, and we cannot be certain that it was definitively caused by rebleeding. Other factors such as hydrocephalus or seizures may have contributed. However, we noted that the distribution of hydrocephalus and seizures at admission was comparable between the RB+ and RB-groups. If our definition had been entirely biased, we would expect to see a discrepancy between the groups at admission in terms of hydrocephalus and seizures, which is not the case in our study. For this reason, we believe our definition remains valid. This might also explain the absence of differences between RB- and RB+ patients regarding the classical risk factors for rebleeding identified in the literature.

Despite these limitations, we still believe that the pragmatic and innovative design of our study, which included several hundred patients, represents a key strength. Our results further reinforce the need for early intervention in the management of aSAH patients.

## Conclusion

The time elapsed since the onset of headache is a significant risk factor for rebleeding in patients admitted for aneurysmal subarachnoid hemorrhage. This finding highlights the critical importance of promptly addressing this delay when planning the aneurysm securing procedure.

## Data Availability

Available on reasonable request.

## Authors’ contribution

Original idea: ED, YL, ALG; Collecting the Data: ED, ALG; Statistical analysis: ED, ALG; First draft of the manuscript: ED; Edition and acceptance of the final version of the manuscript: all authors.

## Fundings

No specific funding was received for this study.

## Conflict of interests

The authors declare no conflicts of interest.

## Notes

### Competing Interest Statement

The authors have declared no competing interest.

### Author Declarations

Comité d’éthique, CHU Rennes, Hôpital Pontchaillou, 2 Rue Henri le Guilloux, 35033 Rennes cedex 9, France;; +33299873553

